# A STUDY TO ASSESS THE KNOWLEDGE AND ATTITUDE REGARDING OPTIONAL VACCINES AND BARRIERS TO USE AMONG MOTHERS OF UNDER FIVE CHILDREN IN KHEDA GUJARAT

**DOI:** 10.1101/2024.12.24.24319589

**Authors:** Sherin Parmar, Shlok Dave, Sneha Vankar, Srusti Selot, Sejal Dabhi, Kailash Nagar

**Author notes:** **Corresponding Author:** Mr. Kailash Nagar (Email Id), Researcher ID: AAM-6294-2021, **Address:** Dinsha Patel College of Nursing, Behind Hyundai Showroom, College road, Nadiad, Kheda District, Gujarat- 387001.

## Abstract

**Introduction:** Vaccination is a straightforward and reliable method to safeguard oneself from dangerous diseases before encountering them. Vaccination remains one of the most effective public health interventions in human history, significantly reducing the burden of infectious diseases and saving countless lives worldwide.

**Aim:** To determine knowledge and attitude regarding optional vaccines and barriers to use among mothers of under five children in Kheda district

**Methodology:** The non-experimental Descriptive Survey Research design used for this study. The study was conducted on 384 mothers of under five children from selected areas of Kheda district by systematic random sampling technique. A knowledge questionnaire tool, Likert attitude scale and barrier questionnaire was used for data collection.

**Result:** 240 (63%) of mothers have low knowledge, 123 (32%) have moderate knowledge, and 21 (5%) have good knowledge regarding optional vaccination. When 1 (0.3%) mother have Unfavourable attitude, 275 (71.6%) have moderate attitude, and 108 (28.1%) have favourable attitude towards optional vaccination. The mean score of attitude is 8.7083. The Correlation-Coefficient (r) of knowledge and attitude is 0.76.

**Conclusion:** The study highlights the disparity between knowledge and attitude regarding optional vaccination among mothers of under five children. Although the majority lack adequate knowledge, a considerable number still maintain a moderate attitude. This emphasizes the need for targeted interventions to improve understanding and promote positive attitudes toward optional vaccination.

## INTRODUCTION

Vaccination is a straightforward and reliable method to safeguard oneself from dangerous diseases before encountering them. By leveraging the body’s innate defences, vaccines induce resistance to particular infections and enhance the immune system. They work by teaching the immune system to generate antibodies, similar to its response when encountering an actual disease. Importantly, vaccines contain only inactive or weakened versions of pathogens such as viruses/subunits, bacteria, or non-infectious RNA, ensuring they don’t cause illness or expose individuals to its actual or potential complications.

The term “vaccine” originates from the Latin word “vaccines,” meaning “from the cows.” Edward Jenner, a pioneering physician and scientist, conducted the first documented scientific attempt to prevent smallpox in 1796. While he didn’t create the method, his systematic study established its effectiveness, earning him recognition as the father of vaccines for his rigorous scientific approach regarding vaccination.

Jenner’s method entailed extracting material from a blister of a person infected with cowpox and introducing it into another person’s skin, known as **arm-to-arm inoculation**. However, advancements in scientific understanding by the end of 1940s enabled the large-scale production of vaccines, marking the beginning of significant efforts in disease control or prevention.

At the start of the 20th century, several vaccines were introduced for routine use, including those for pertussis in 1914, diphtheria in 1926, and tetanus in 1938. In 1948, these three vaccines were combined to create the DTP vaccine.

Vaccination remains one of the most effective public health interventions in human history, significantly reducing the burden of infectious diseases and saving countless lives worldwide. However, the issue of optional vaccination has sparked considerable debate in recent years, with discussions revolving around personal choice, public health responsibilities, and the balance between individual freedoms and community protection.

Optional vaccines, also known as elective or non-mandatory vaccines, are a topic of considerable debate and discussion in public health and policy circles. Unlike mandatory vaccines that are required by law for certain groups or for attending school, optional vaccines are recommended by health authorities but are not compulsory. This essay explores the concept of optional vaccines, their significance, controversies surrounding them, and their implications for public health.

Optional vaccines encompass a range of immunizations that are not universally mandated but are recommended based on individual risk factors, age, lifestyle, or occupation. These vaccines often target diseases that may not pose a significant public health threat in all circumstances but could be detrimental to specific populations. Examples include vaccines for diseases like HPV (Human Papillomavirus), meningococcal disease, and hepatitis A and B.

Moreover, optional vaccines can play a crucial role in protecting vulnerable populations. For instance, vaccines against diseases like influenza are recommended for certain groups such as the elderly and those with compromised immune systems. By choosing to vaccinate, individuals not only protect themselves but also contribute to herd immunity, reducing the overall transmission of infectious diseases within communities.

## OBJECTIVE

- To assess the knowledge regarding the optional vaccines and barriers to use it among the mothers of under five children.
- To assess the attitude regarding optional vaccines and barriers for use among mothers of under five children.
- To assess the significant association between level of knowledge, attitude score and their selected demographic variables.

## METHODS

This descriptive study was conducted after institutional ethical committee approval. A pre-validated knowledge questionnaire tool, Likert attitude scale And Barrier Questionnaire regarding organ donation was circulated to 384 Mothers of under-five children from selected village of Kheda district by used non probability purposive sampling technique from 25/06/2024 to 01/07/2024. Written informed consent was obtained from all mothers of under-five children, along with assent form.

For knowledge questionnaire tool regarding optional vaccination, the total minimum score is 0 and total maximum score is 18. The cut off score and category were calculated as 14-18 indicate good knowledge, 9-13 indicate average knowledge and below 9 indicate poor knowledge regarding optional vaccination. For Likert attitude scale regarding optional vaccination, the total minimum score is 10 and total maximum score is 30. The cut off score and category were calculated as 23-30 indicate favourable attitude, 15-22 indicate moderate attitude and below 15 indicate unfavorable attitude regarding optional vaccination. The mothers were included from Salun Village, Dist.: Kheda. Data of both components were distributed in percentage based on mother’s age, child’s age, religion, type of family, mother’s education, father’s education family monthly income, source of immunization, the association between knowledge regarding optional vaccination with their selected demographic variables and also between attitudes regarding optional vaccination with their selected demographic variables determined. Correlation between knowledge and attitude regarding optional vaccination is also determined.

### Ethical Considerations

Ethical considerations for this study included obtaining approval from the principal and the institute’s ethical committee. Informed consent was obtained from all participants, who were fully informed that their participation was entirely voluntary.

## RESULTS

### Demographic key findings of the study

Demographic data of 384 mothers were normally distributed. In that, 175 (45.57%) samples were between the age group of 25.1-30 years, 119 (30.98%) samples were between the age group of 1-3 years, 352 (91.66%) samples were Hindu Family, 234(60.94%) of samples were Joint Family, 145 (37.76%) mothers had primary education, 128 (33.37%) fathers had secondary education, 160 (41.67%) families had income less then □10,000, 214 (55.73%) family was provided information regarding vaccination by Health Caregiver and 340 (88.55%) mother did not provided information regarding optional vaccine.

In the distribution and frequency of knowledge regarding optional vaccination among mothers of under-five children, there are 240 (63%) of mothers have low knowledge, 123 (32%) have moderate knowledge, and 21 (5%) have good knowledge regarding optional vaccination. The mean score of knowledge is 8.7083. Where the distribution and frequency of attitude regarding optional vaccination among mothers of under-five children, 1 (0.3%) mother have Unfavorable attitude, 275 (71.6%) have moderate attitude, and 108 (28.1%) have favorable knowledge. The mean score of attitude is 21.1641

Table-2 shows Range, Mean and Standard Deviation for Level of Knowledge and Level of Attitude. Knowledge score has Range of 15.00, Mean is 8.7083 and Standard deviation is 2.91182. Attitude score has Range of 13.00, Mean is 21.1641 and Standard deviation is 2.25795.

**Graph:1.**
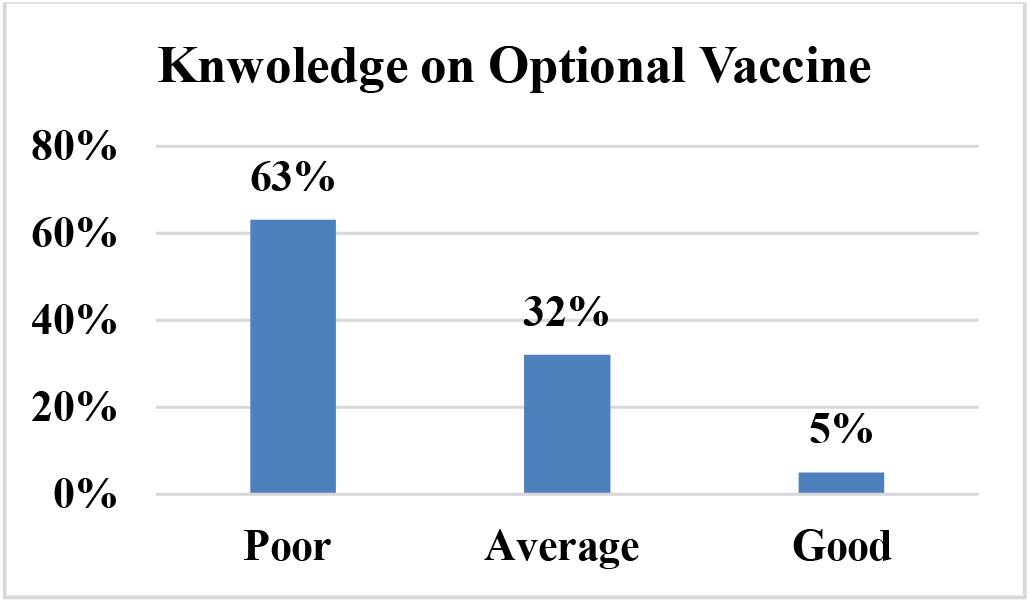
Level of knowledge on optional Vaccine among Mothers.

**Graph:2.**
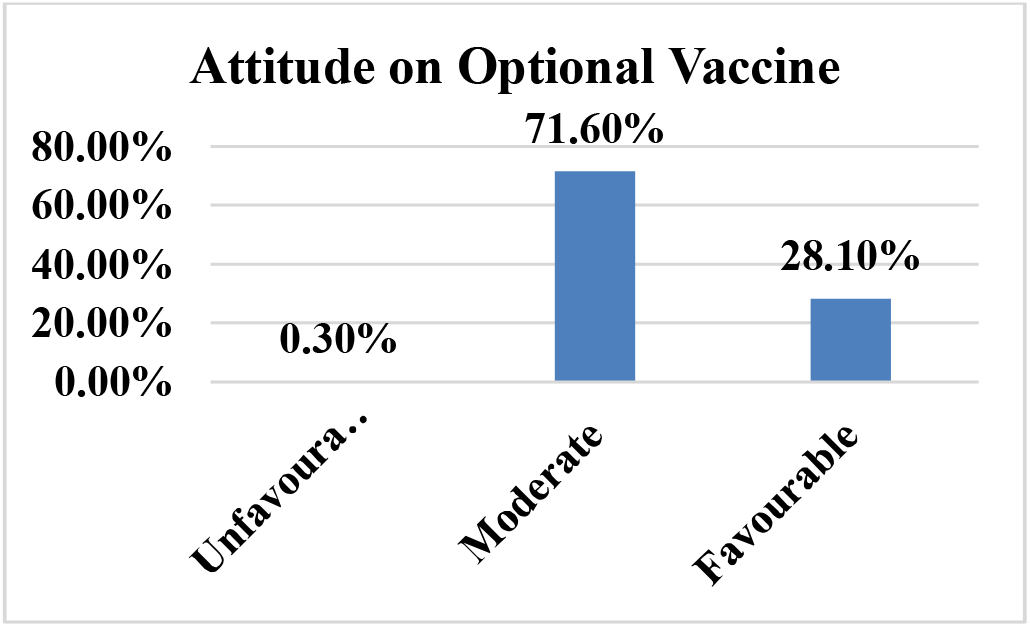
Level of knowledge on optional Vaccine among Mothers.

**Table 1:** Dimorphic variable of the participants (n=384)

| Demographic Variable | Frequency | Percentage |
| --- | --- | --- |
| **Age of Mother:** |  |  |
| 18-25 years | 126 | 32.83% |
| 25.1-30 years | 175 | 45.57% |
| 30.1-35 years | 60 | 15.62% |
| Above 35 | 23 | 5.98% |
| **Age of Child** |  |  |
| 0-6 months | 45 | 11.72% |
| 6 months- 1year | 110 | 28.65% |
| 1-3 years | 119 | 30.98% |
| 3-5 years | 110 | 28.65% |
| **Religion** |  |  |
| Hindu | 352 | 91.66% |
| Christian | 25 | 6.51% |
| Muslim | 7 | 1.83% |
| Other | 0 | 0% |
| **Type of Family** |  |  |
| Nuclear | 150 | 39.06% |
| Joint | 234 | 60.94% |
| Other | 0 | 0% |
| **Education of Mother** |  |  |
| No formal Education | 48 | 12.50% |
| Primary Education | 145 | 37.76% |
| Secondary Education | 109 | 28.39% |
| Graduate & above | 82 | 21.35% |
| **Education of Father** |  |  |
| No formal Education | 24 | 6.29% |
| Primary Education | 113 | 29.35% |
| Secondary Education | 128 | 33.37% |
| Graduate & above | 119 | 30.99% |
| **Monthly Family Income** |  |  |
| < ₹10,000 | 160 | 41.67% |
| ₹10,001- ₹20,000 | 103 | 26.82% |
| ₹20,001- ₹30,000 | 71 | 18.48% |
| > ₹30,000 | 50 | 13.03% |
| **Source of Immunization information** |  |  |
| Healthcare Provider | 214 | 55.73% |
| Family/Friends | 27 | 7.03% |
| Internet | 34 | 8.85% |
| Television/Radio | 6 | 1.56% |
| Govt. Health Campaign | 81 | 21.10% |
| Other | 22 | 5.73% |
| **Has your child received any optional vaccine?** |  |  |
| Yes | 44 | 11.45% |
| No | 340 | 88.55% |

**Table-2:** Frequency and distribution of level of knowledge and attitude regarding optional vaccines among mothers of under five children.

| Knowledge |  | Attitude |  |
| --- | --- | --- | --- |
| Poor | 240 (63%) | Unfavorable | 1 (0.3%) |
| Average | 123 (32%) |  | 275 (71.6%) |
| Good | 21 (5%) | Favorable | 108 (28.1%) |

**Table-2:** Range, Mean and Standard Deviation for Level of Knowledge and Level of Attitude.

| Mean and SD | Range | Mean | SD |
| --- | --- | --- | --- |
| **Knowledge Score** | 15 | 8.70 | 2.91 |
| **Attitude Score** | 13 | 21.1 | 2.25 |

Table 3 illustrates the correlation between knowledge and attitude regarding optional vaccines among 384 mothers of under-five children. The analysis reveals a strong positive correlation (r = 0.76) between the two variables, which is statistically significant. This indicates that higher knowledge about optional vaccines among mothers is associated with a more positive attitude towards them, emphasizing the need for effective educational programs to improve awareness and foster favorable attitudes.

**Table-3:** Correlation between Knowledge and Attitude regarding optional vaccines among mothers of under five children. (n=384)

| Correlation | R score |
| --- | --- |
| **Knowledge** | **0.76**<br>**S** |
| **Attitude** |  |

**Table-4:** Barriers regarding optional vaccines among mothers of under five children.

| QUESTIONS | Yes |  | No |  |
| --- | --- | --- | --- | --- |
|  | F | % | F | % |
| Are you adequately aware about the optional vaccine that are available? | 256 | 67% | 128 | 33% |
| Does the cost of optional vaccines affect your decision to get them to your child? | 224 | 58% | 160 | 42% |
| Do you feel that optional vaccines are safe for your child? | 263 | 68% | 121 | 32% |
| Are there any issues related to transportation to health care facilities for vaccinating your child? | 199 | 52% | 185 | 48% |
| Are you adequately informed by your health care professionals about the benefits of optional vaccines? | 195 | 51% | 189 | 49% |
| Do you worry about the pain that your child might experience during vaccination process? | 251 | 65% | 133 | 35% |
| Do you think that your local health care services should provide optional vaccines free of cost? | 263 | 68% | 121 | 32% |
| Do you require a time-to-time reminder for vaccinating your child? | 221 | 58% | 163 | 42% |
| Are there any social stigmas within your community that affects your decision to vaccinate your child? | 197 | 51% | 187 | 49% |

**Table-5:** Association between level of Knowledge and Demographic variables of the study participants.

| Demographic Variable | $\chi^2$ | df | P-Value |
| --- | --- | --- | --- |
| **Age of Mother:** | **13.613** | **6** | **0.034 (S)** |
| 18-25 years |  |  |  |
| 25.1-30 years |  |  |  |
| 30.1-35 years |  |  |  |
| Above 35 |  |  |  |
| **Age of Child** | **12.718** | **6** | **0.048 S** |
| 0-6 months |  |  |  |
| 6 months- 1year |  |  |  |
| 1-3 years |  |  |  |
| 3-5 years |  |  |  |
| **Religion** | 0.813 | 4 | 0.937 NS |
| Hindu |  |  |  |
| Christian |  |  |  |
| Muslim |  |  |  |
| Other |  |  |  |
| **Type of Family** | 5.758 | 2 | 0.056 NS |
| Nuclear |  |  |  |
| Joint |  |  |  |
| Other |  |  |  |
| **Education of Mother** | **35.351** | **6** | **0.000 S** |
| No formal Education |  |  |  |
| Primary Education |  |  |  |
| Secondary Education |  |  |  |
| Graduate & above |  |  |  |
| **Education of Father** | **22.240** | **6** | **0.001 S** |
| No formal Education |  |  |  |
| Primary Education |  |  |  |
| Secondary Education |  |  |  |
| Graduate & above |  |  |  |
| **Monthly Family Income** | **31.190** | **6** | **0.000 S** |
| < ₹10,000 |  |  |  |
| ₹10,001- ₹20,000 |  |  |  |
| ₹20,001- ₹30,000 |  |  |  |
| > ₹30,000 |  |  |  |
| **Source of Immunization information** | **21.513** | **10** | **0.018 S** |
| Healthcare Provider |  |  |  |
| Family/Friends |  |  |  |
| Internet |  |  |  |
| Television/Radio |  |  |  |
| Govt. Health Campaign |  |  |  |
| Other |  |  |  |

**Table-6:** Association between level of Attitude and Demographic variables of the study participants.

| Demographic Variable | $\chi^2$ | df | P-Value |
| --- | --- | --- | --- |
| **Age of Mother:** | 9.102 | 6 | 0.168 NS |
| 18-25 years |  |  |  |
| 25.1-30 years |  |  |  |
| 30.1-35 years |  |  |  |
| Above 35 |  |  |  |
| **Age of Child** | 5.773 | 6 | 0.449 NS |
| 0-6 months |  |  |  |
| 6 months- 1year |  |  |  |
| 1-3 years |  |  |  |
| 3-5 years |  |  |  |
| **Religion** | 8.534 | 4 | 0.074 NS |
| Hindu |  |  |  |
| Christian |  |  |  |
| Muslim |  |  |  |
| Other |  |  |  |
| **Type of Family** | 2.057 | 2 | 0.358 NS |
| Nuclear |  |  |  |
| Joint |  |  |  |
| Other |  |  |  |
| **Education of Mother** | 14.357 | 6 | 0.026 S |
| No formal Education |  |  |  |
| Primary Education |  |  |  |
| Secondary Education |  |  |  |
| Graduate & above |  |  |  |
| **Education of Father** | 8.714 | 6 | 0.190 NS |
| No formal Education |  |  |  |
| Primary Education |  |  |  |
| Secondary Education |  |  |  |
| Graduate & above |  |  |  |
| **Monthly Family Income** | **19.362** | **6** | **0.004 S** |
| < ₹10,000 |  |  |  |
| ₹10,001- ₹20,000 |  |  |  |
| ₹20,001- ₹30,000 |  |  |  |
| > ₹30,000 |  |  |  |
| **Source of Immunization information** | 2.620 | 10 | 0.989 NS |
| Healthcare Provider |  |  |  |
| Family/Friends |  |  |  |
| Internet |  |  |  |
| Television/Radio |  |  |  |
| Govt. Health Campaign |  |  |  |
| Other |  |  |  |

## CONCLUSION

The study revealed a significant gap in knowledge regarding optional vaccines among mothers of under-five children, with 63% of mothers exhibiting poor knowledge, while only 5% demonstrated good knowledge. Despite this, a considerable number of mothers (71.6%) exhibited a moderate attitude toward optional vaccination, with 28.1% showing a favorable attitude. The strong positive correlation (r = 0.76) between knowledge and attitude highlights the need for targeted educational programs to bridge this knowledge gap and foster more positive attitudes toward optional vaccines. Barriers such as cost, lack of adequate information from healthcare providers, concerns about vaccine safety, transportation issues, and social stigma were identified as significant challenges in the uptake of optional vaccines. The study underscores the importance of implementing cost-effective, accessible, and community-centered vaccination awareness campaigns to address these barriers and enhance the overall immunization coverage.

These findings emphasize the crucial role of healthcare providers and public health policies in promoting awareness and ensuring that mothers are well-informed about the benefits and availability of optional vaccines, ultimately contributing to improved child health outcomes.

## DISCUSSION

The study revealed significant gaps in knowledge and varying attitudes among mothers of under-five children regarding optional vaccines, with the majority demonstrating poor knowledge and moderate attitudes. A strong positive correlation between knowledge and attitude highlights the need for targeted educational interventions to improve awareness and foster favorable perceptions. Key barriers to vaccine uptake included high costs, lack of awareness, safety concerns, and logistical challenges, consistent with findings from similar studies. These barriers underscore the importance of implementing affordable vaccination programs, enhancing healthcare communication, and increasing accessibility through community-based initiatives. While the study provides valuable insights, its cross-sectional design and focus on a specific region may limit generalizability. Overall, addressing these challenges through strategic public health efforts can improve vaccine uptake and contribute to better child health outcomes.

## Data Availability

yes authors are ready to provide all the details whenever it require

## Statement of Informed Consent

The informed consent form was taken from the postnatal mothers prior to the data collection of the study.

## REFERENCES

1. World Health Organization (Internet) [cited 2024], available from, https://www.who.int/news-room/feature-stories/detail/the-race-for-a-covid-19-vaccine-explained

2. Geneva: WHO; 2012. Mar, [accessed on May 30, 2012]. World Health Organization. Global immunization data 2011. https://www.who.int/hpvcentre/Global_Immunization_Data.pdf.

3. Saraswathi KN, Lissa J. A study to assess the knowledge on selected optional vaccines among mothers of under-five children in selected immunization centers at Mysore with a view to develop an information booklet. Asian J Nurs Educ Res. 2014;4(4):513–515. Available from,https://ajner.com/AbstractView.aspx?PID=2014-4-4-24

4. Hazaratali Panari, Anuchithra. Study on Immunization among the Mothers of Under-five Children, Halaga Village, Belgaum, Karnataka. Asian J. Nur. Edu. and Research. 2016; 6(2): 191–198. doi: 10.5958/2349-2996.2016.00035.5 Available on: https://ajner.com/AbstractView.aspx?PID=2016-6-2-8

5. Christian S, Ravindran HN. A study to assess the knowledge on Hib vaccination among the mothers of neonates in Dhiraj Hospital. Int J Sci Res. 2015;4(5):218–221.Available from: https://www.worldwidejournals.com/international-journal-of-scientific-research-(IJSR)/recent_issues_pdf/2015/May/May_2015_1492855573__218.pdf

6. Fredy S. Knowledge, attitude, and practice regarding optional vaccination among mothers of under-five children. Int J Sci Acad Res. 2019;1(4):12–20. Available from: https://www.ijsar.in/Admin/pdf/knowledge-attitude-and-practice-regarding-optional-vaccination-among-mothers-of-under-five-children-.pdf

7. Ambike D, Tambade V, Poker F, Ahmed K. Parental knowledge on the optional vaccines and the barriers in their use: A rural hospital-based study. Indian J Child Health. 2016;4:88–9.

8. Naik, J.D., Jain, S., Babar, S.D., Radhey, B.K., Kamble, G. and Gajbhijiye, R. 2016. Awareness of Measles among mothers of under-five children Attending UHC Immunization Clinic of Government Medical College

9. Gul S, Khalil E. Mothers’ knowledge and practices related to immunization at a tertiary care hospital in Karachi, Pakistan. Int J Adv Med. 2016 Aug;3(3):656–61. Available from: http://www.ijmedicine.com

